# Cremation based estimates suggest significant under- and delayed reporting of COVID-19 epidemic data in Wuhan and China

**DOI:** 10.1101/2020.05.28.20116012

**Authors:** Mai He, Li Li, Louis P. Dehner, Lucia F. Dunn

## Abstract

**Background:** China’s COVID-19 statistics fall outside of recognized and accepted medical norms. Here we estimated the incidence, death and starting time of the COVID-19 outbreak in Wuhan and China based on cremation related information.

**Methods:** Data sources included literature on COVID-19 in China, official Chinese government figures, state-run and non state-run media reports. Our estimates are based on investigative media reports of crematory operations in Wuhan, which is considered as a common data end point to life. A range of estimates is presented by an exponential growth rate model from lockdown (Jan 23,2020) until the intervention started to show effects, which was estimated 14.5 days after lockdown.

**Results:** For the cumulative infections and total deaths, under different assumptions of case fatality rates (from 2.5% to 10%) and doubling time 6.4 days, the estimates projected on February 7, 2020 in Wuhan range from 305,000 to 1,272,000 for infections and from 6,811 to 7,223 for deaths - on the order of at least 10 times the official figures (13,603 and 545). The implied starting time of the outbreak is October 2019. The estimates of cumulative deaths, based on both funeral urns distribution and continuous full capacity operation of cremation services up to March 23, 2020, give results around 36,000, more than 10 times of the official death toll of 2,524.

**Conclusions:** Our study indicates a significant under-reporting in Chinese official data on the COVID-19 epidemic in Wuhan in early February, the critical time for response to the COVID-19 pandemic.

## BACKGROUND

The spread of the coronavirus (COVID-19) has evolved into a global public health crisis affecting all aspects of individual, societal and economic activity. On December 31, 2019, the Wuhan City Health Commission announced that recent cases of pneumonia of unknown etiology seen in some hospitals were related to the Wet Market and 27 cases were found by the time. This official announcement conflicted with at least two official numbers and one media report which were 45, 104 and 266 cases.^1–3^ Reports from the official Chinese state-run media on February 2, 2020 showing large numbers of possible coronavirus victims who were not treated within the medical establishment and hence may have fallen outside of government statistics, have led many to believe there may be serious gaps in our understanding of the outbreak based on what can be determined from this official government data.^4^

As the epicenter of the COVID-19 initial outbreak, the epidemiological information from Wuhan affects the response and preparation of other parts of China and rest of the world. It is therefore important to attempt to accurately assess the actual number of cases and gain some insights from this deduction to the time of the first cases.

The purpose of this report is to investigate the epidemiological information of the early phase of the COVID-19 outbreak in Wuhan, after lockdown on January 23, 2019, based on official Chinese government figures, published literature and media reports, focusing on cremation related data. The estimates of incidence and deaths are higher by a magnitude of at least 10 times the official figures. Our estimates are further supported by others who were also investigating the situation and recent updates of funeral urn distribution in Wuhan. The potential impact of this discrepancy is critical for both the medical projection of needs and for policy decisions relating to public health.

## METHODS

### Data Sources

The official Chinese government figures include national and local Wuhan data^5^ and state-run media reports. Since there is no medical literature on “COVID-19, cremation” in China, we searched on the internet for media reports in Chinese by “新型冠状肺炎” (novel coronavirus pneumonia) and “Wuhan” on the operation of crematory facilities in Wuhan as widely reported by established news organizations both inside and outside of China.^6–12^

## Cremation Data

Daily cremation due to the COVID-19 outbreak is defined and estimated by total number of estimated cremations in the time period of this study minus pre-outbreak average daily cremations.

Detailed information on crematory facility and operation is listed in Appendix 1, but also summarized in Table 1.^6,7^ Briefly, there are eight crematories in Wuhan, which under normal circumstances would operate about 4 hours per day. Before the outbreak, cremation mainly happened in the morning, according to Chinese rituals. Starting on or before January 25, 2020, these were observed to be operating at or close to around-the-clock or 24 hours daily.^6–12^ This would put the current operating rate at about six times normal. Normal deaths per day can be estimated as 136 based on an annual mortality rate of 0.00551 in a population of approximately 9 million (Wuhan government data).^13^ With the regular procedure, the additional 20 hours of daily operation imply additional deaths of 5 (20/4) x 136 = 680 per day above normal, if the services are utilized with full capacity. However, it was estimated that the maximal capacity of cremation could be up to 2,100 bodies per day (see appendix 1).

**Table 1.** Crematory Facilities and Operation in Wuhan, China.

|  | **Before lockdown on January 23, 2020** | **From January 25 to February 19, 2020** | **After February 19, 2020** |
| --- | --- | --- | --- |
| **Crematory facilities** | 86 cremation furnaces* | 86 cremation furnaces* | 86 cremation furnaces*<br>40 mobile crematory stations |
| **Crematory operation** | Four hours per day | 24 hours per day | 24 hours per day |
| **Crematory capacity** | Approximately 136 per day | 816 per day (full capacity)<br>2,064 (maximum capacity) ** | 2,064 and 200 tons per day (5 tons “medical waste”<br>including “animal bodies” per mobile crematory<br>station, total capacity 5x40 = 200 tons) |

\* The furnace information is for seven funeral houses, Hankou, Wuchang, Qingshan, Caidian District, JiangXia Distric, Huangpi District, Xinzhou District. Muslim Funeral House is not included in the table above, since the information there is scarce.
\*\* The maximum capacity is calculated for the seven funeral houses, assuming non-stop cremation, and simplified procedure, since no farewell ceremony was allowed during lockdown period.

This estimate was supported by media reports of support of additional crematory staff from other cities and provinces being brought into Wuhan.^14,15^ Since February 19, 40 mobile crematory stations were also sent to Wuhan for the increasing need of cremation capacity.^16,17^

### Estimation

We used an exponential model to estimate the cumulative infections and deaths in Wuhan.

### Assumptions

I. The median time of the incubation period for COVID-19 is 4.5 days from exposure to symptom onset.^18^ The median time from the symptom onset to death is 14 days. ^19^ Assuming it takes half a day from death to cremation, this implies that most of the deaths which happened on and before February 11, 2020 were not impacted by any countermeasures in effect.
II. In this study, different assumptions of epidemic doubling time were applied. Wu et al. statistically inferred case counts in Wuhan by internationally exported cases as of January 25 and estimated doubling time as 6.4 days with 95% confidence interval (5.8, 7.1).^20^ Rodriguez et al. analyzed Chinese official daily cumulative cases from January 20 to February 9, 2020, and derived the doubling time for each province ranging from 1.42 to 3.05 days, with Hubei (where Wuhan is located) as 2.54 with confidence interval (2.44, 2.64), based on an exponential model. With no countermeasure in effect, we assumed doubling time was also 2.54 days before January 20 in this scenario.^21^
III. Outcomes are either survival with probability (1 − *d*) where d is the case fatality rate; or death after 14 days with probability *d*.
IV. To extrapolate from the Wuhan cremation data to the total number of infected cases, alternative assumptions of 2.5%, 5%, and 10% were utilized for the case fatality rate.
V. Though media reported that Wuhan cremation service operated around the clock since January 25, 2020,^6–12^ we assumed no full capacity was utilized until two weeks later, on February 7, 2020. This assumption will be further discussed below.

### The Model

Estimates of the total cases were calculated from an exponential growth model. At time *t*, where *t* is the number of days since the occurrence of the initial case, the quantities of interest are *N*(*t*), the cumulative number of cases, *n*(*t*), the daily cases, *D*(*t*), the cumulative number of deaths, *d*(*t*), the daily deaths, and the case fatality rate *d. N*(*t*) = *c*(*eλ^t^* − 1) + 1, so that *N*(*0*) *=* 1, where *c* is a constant and the parameter *λ* is set to match the reported doubling time. *n*(*t*) ≅ (1 − *e*^−^*^λ^*) *N*(*t*) and *D*(*t*) *= d e^−^*^14.5^ *^λ^N*(*t*). Matching the cumulative totals N(t) to the model where each existing case infects *R*_0_ (the basic reproductive number) new cases on average after s days (the serial interval) gives *c* = *R*_0_/*R*_0_ − 1). Wu et al. estimates 2.7 for *R*_0_.^20^ Exponential growth model was not applied for projections of N(t) for Wuhan beyond the date February 7, 2020, due to the declining number of susceptible individuals and plausible effectiveness of city lockdown.

The projections of *N*(*t*) for Wuhan using doubling time 2.54 days approach half million on 44^th^ day, that the exponential growth model is unreliable. From the 45^th^ day, we take into account the declining number of susceptible individuals, and the model was modified (as in the SIR simulation model) so that the relationship *n*(*t*) = *e^λ^n*(*t −* 1) becomes 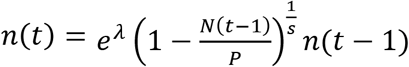 where *P* is the population size of Wuhan after lock down.

The 95% confidence intervals presented in Table 2 are based on the confidence interval for the doubling rate as presented. Confidence intervals for the estimated starting dates are not provided because they depend on estimates of both *λ* and *R*_0_ and also on the randomness in the transmission of the infection in its very early stages.

**Table 2.** Official COVID-19 Epidemic Data in Wuhan and China from January 23 to April 17, 2020.

| **Date** | **Cumulative confirmed infections in Wuhan** | **Cumulative deaths in Wuhan** | **Fatality rate in Wuhan** | **Cumulative confirmed infections in China** | **Cumulative deaths in China** | **Fatality rate in China** | **Confirmed cases W/C** | **Deaths W/C** |
| --- | --- | --- | --- | --- | --- | --- | --- | --- |
| **23-Jan**<br>(lockdown) | 495 |  |  |  |  |  |  |  |
| 25-Jan | 618 |  |  |  |  |  |  |  |
| 4-Feb | 8351 | 362 | 4.33% | 24324 | 490 | 2.01% | 34.3% | 73.9% |
| 5-Feb | 10117 | 417 | 4.12% | 28018 | 563 | 2.01% | 36.1% | 74.1% |
| 6-Feb | 11618 | 478 | 4.11% | 31161 | 636 | 2.04% | 37.3% | 75.2% |
| 7-Feb | 13603 | 545 | 4.01% | 34546 | 722 | 2.09% | 39.4% | 75.5% |
| 8-Feb | 14982 | 608 | 4.00% | 37198 | 811 | 2.18% | 40.3% | 75.0% |
| 9-Feb | 16902 | 681 | 4.03% | 40171 | 908 | 2.26% | 42.1% | 75.0% |
| 10-Feb | 18454 | 748 | 4.05% | 42638 | 1016 | 2.38% | 43.3% | 73.6% |
| 11-Feb | 19558 | 820 | 4.19% | 44653 | 1113 | 2.49% | 43.8% | 73.7% |
| **12-Feb** | 32994 | 1036 | 3.14% | 59804 | 1367 | 2.29% | 55.2% | 75.8% |
| 13-Feb | 35991 | 1016 | 2.82% | 63851 | 1380 | 2.16% | 56.4% | 73.6% |
| **23-March** | 50006 | 2524 | 5.05% | 81171 | 3277 | 4.04% | 61.6% | 77.0% |
| 11-April | 50008 | 2577 | 5.15% | 82052 | 3339 | 4.07% | 60.9% | 77.2% |
| **17-April** | 50,333 | 3,859 | 7.69% | 82,692 | 4,632 | 5.60% | 60.9% | 83.3% |

## RESULTS

Wuhan lockdown started on January 23, 2020. This study chose February 7 (two weeks after the announcement of 24-hour-operation of cremation services), and February 12 (20 days after lockdown) as the projected evaluation days. Retrospectively, February 12 was the day when party leadership changes were undertaken in Wuhan and Hubei Province.^22^

### I. Chinese official data for Wuhan and China

The Chinese government’s official COVID-19 cumulative confirmed diagnoses and deaths data, from late January up to mid-February, and selected representative dates later, are recorded in Table 2 for Wuhan and the whole of China.^23^ Daily ratios of confirmed diagnosis and death between Wuhan and China as whole were calculated and listed in Table 2. While the increase was relatively stable throughout the early period shown, there was a marked addition of confirmed cases on February 12, 2020, with 13,436 new cases reported in Wuhan and a total of 15,151 (including Wuhan’s data) of new cases in all of China, amounting to cumulative confirmed diagnoses of 32,994 cases in Wuhan and 59,804 cases in China as a whole, and 1,036 cases of cumulative deaths in Wuhan and 1,367 deaths in China. On February 12, 2020, Wuhan’s cases represented 55.2% and 75.8% of China’s cases of cumulative confirmed diagnoses and deaths, respectively. The case fatality rate due to COVID-19 ranged from 3 to 4% in Wuhan during this period, and ultimately 5% by April 11, 2020 (Table 1).

On April 17, Wuhan City announced an adjustment of its death toll by additional 1,290, and of confirmed diagnosis by 325 cases, making a total of confirmed diagnosis of 50,333 cases, deaths of 3,869 cases, and crude case fatality rate of 7.69%.^24,25^

### II. Estimates of COVID-19 related deaths and cumulative infections in Wuhan

a. *Simple linear calculation* As a benchmark comparison, we have provided simple linear calculations with constant daily deaths. We assume that cremation service did not reached 100% utilization until February 7, and the utilization was 80% between January 25 to February 6, 2020. The results of these estimates are listed in Table 3. Cumulative deaths are 9,384, for the nineteen days (January 25 to February 12, 2020) only. Based on the official crude case fatality rate of 3.14% (February 12, 2020, Table 1), the estimated cumulative infection was 298,854 within nineteen days (Table 3).
b. Exponential growth model Based on the cremation data inference that COVID-19 related deaths reached 680 on February 7, 2020, estimation of infection on February 7 is calculated. With 6.4 day doubling rate,^20^ and under assumption of the crude fatality rates of 2.5%, 5%, and 10%, cumulative infections in Wuhan are 1,272,400, 650,900 and 305,000 respectively. Cumulative deaths are 6,946, 7,223 and 6,811 respectively. The implied start dates for the outbreak are October 4, 2019, October 11, 2019 and October 17, 2019, respectively. Estimates based of a 2.54 day doubling rate are also presented with estimated confidence intervals in Table 3. Since this doubling time was derived using Chinese official data, we applied the official 3.14% fatality rate in the calculation. The total cases are projected to be 2.23 million with 95% confidence interval (2.12 million, 2.38 million) on February 7, 2020.
c. Estimates based on funeral urn distribution No one was allowed to pick up urns during the lockdown. Beginning on March 23, Wuhan residents were allowed to collect crematory urns, with the target date as April 4 to complete the distribution of the backlog. Estimates based urn collection are listed in Table 4.

**Table 3.**
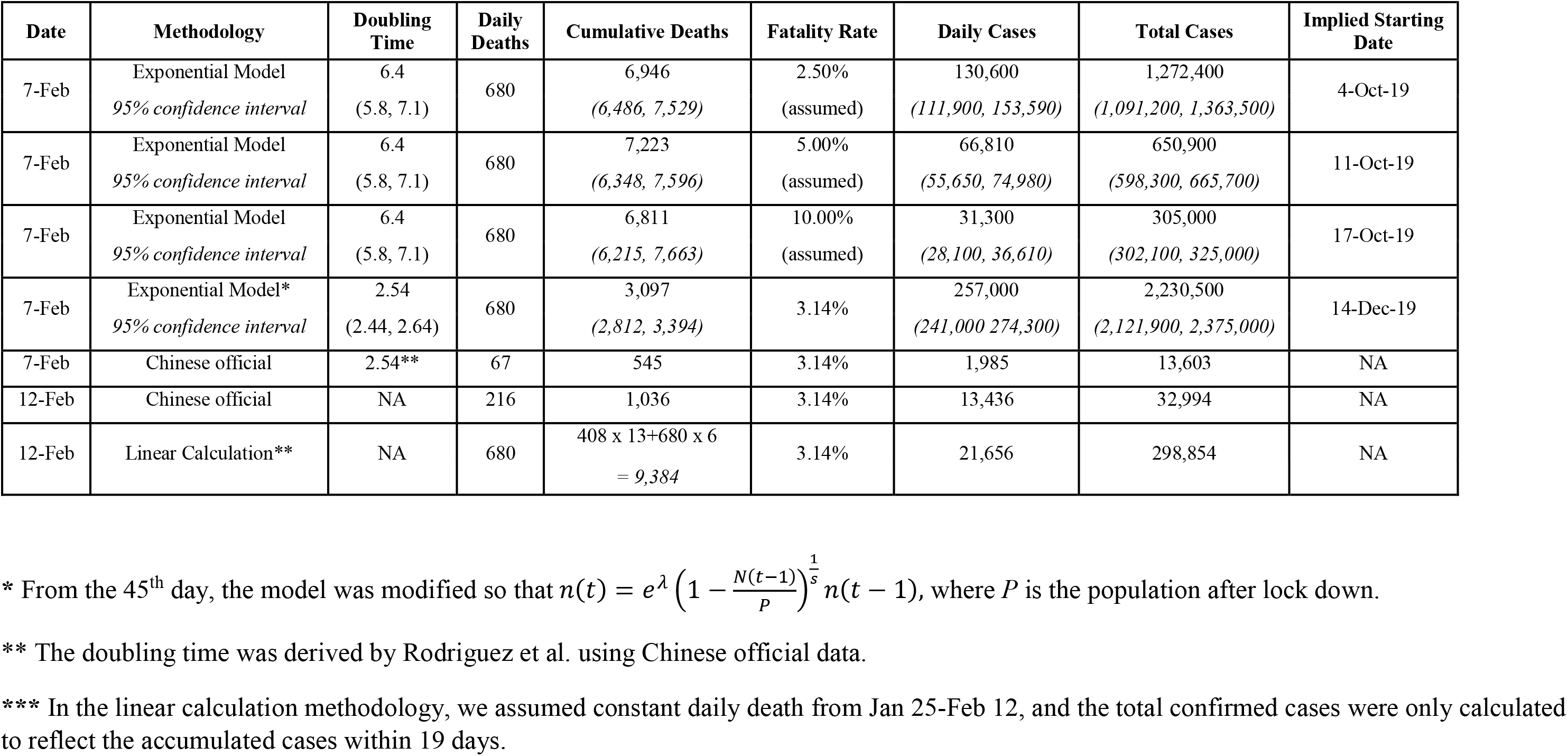
Estimates of COVID-19 Outbreak in Wuhan, China for Feb 7 and Feb 12, 2020.

| Date | Methodology | Doubling Time | Daily Deaths | Cumulative Deaths | Fatality Rate | Daily Cases | Total Cases | Implied Starting Date |
| --- | --- | --- | --- | --- | --- | --- | --- | --- |
| 7-Feb | Exponential Model<br>*95% confidence interval* | 6.4<br>(5.8, 7.1) | 680 | 6,946<br>(6,486, 7,529) | 2.50%<br>(assumed) | 130,600<br>(111,900, 153,590) | 1,272,400<br>(1,091,200, 1,363,500) | 4-Oct-19 |
| 7-Feb | Exponential Model<br>*95% confidence interval* | 6.4<br>(5.8, 7.1) | 680 | 7,223<br>(6,348, 7,596) | 5.00%<br>(assumed) | 66,810<br>(55,650, 74,980) | 650,900<br>(598,300, 665,700) | 11-Oct-19 |
| 7-Feb | Exponential Model<br>*95% confidence interval* | 6.4<br>(5.8, 7.1) | 680 | 6,811<br>(6,215, 7,663) | 10.00%<br>(assumed) | 31,300<br>(28,100, 36,610) | 305,000<br>(302,100, 325,000) | 17-Oct-19 |
| 7-Feb | Exponential Model*<br>*95% confidence interval* | 2.54<br>(2.44, 2.64) | 680 | 3,097<br>(2,812, 3,394) | 3.14% | 257,000<br>(241,000, 274,300) | 2,230,500<br>(2,121,900, 2,375,000) | 14-Dec-19 |
| 7-Feb | Chinese official | 2.54** | 67 | 545 | 3.14% | 1,985 | 13,603 | NA |
| 12-Feb | Chinese official | NA | 216 | 1,036 | 3.14% | 13,436 | 32,994 | NA |
| 12-Feb | Linear Calculation** | NA | 680 | 408 x 13 + 680 x 6<br>= 9,384 | 3.14% | 21,656 | 298,854 | NA |

\* From the 45<sup>th</sup> day, the model was modified so that $n(t) = e^{\lambda} \left(1 - \frac{N(t-1)}{P}\right)^{\frac{1}{s}} n(t-1)$ , where $P$ is the population after lock down.
\*\* The doubling time was derived by Rodriguez et al. using Chinese official data.
\*\*\* In the linear calculation methodology, we assumed constant daily death from Jan 25-Feb 12, and the total confirmed cases were only calculated to reflect the accumulated cases within 19 days.

**Table 4.**
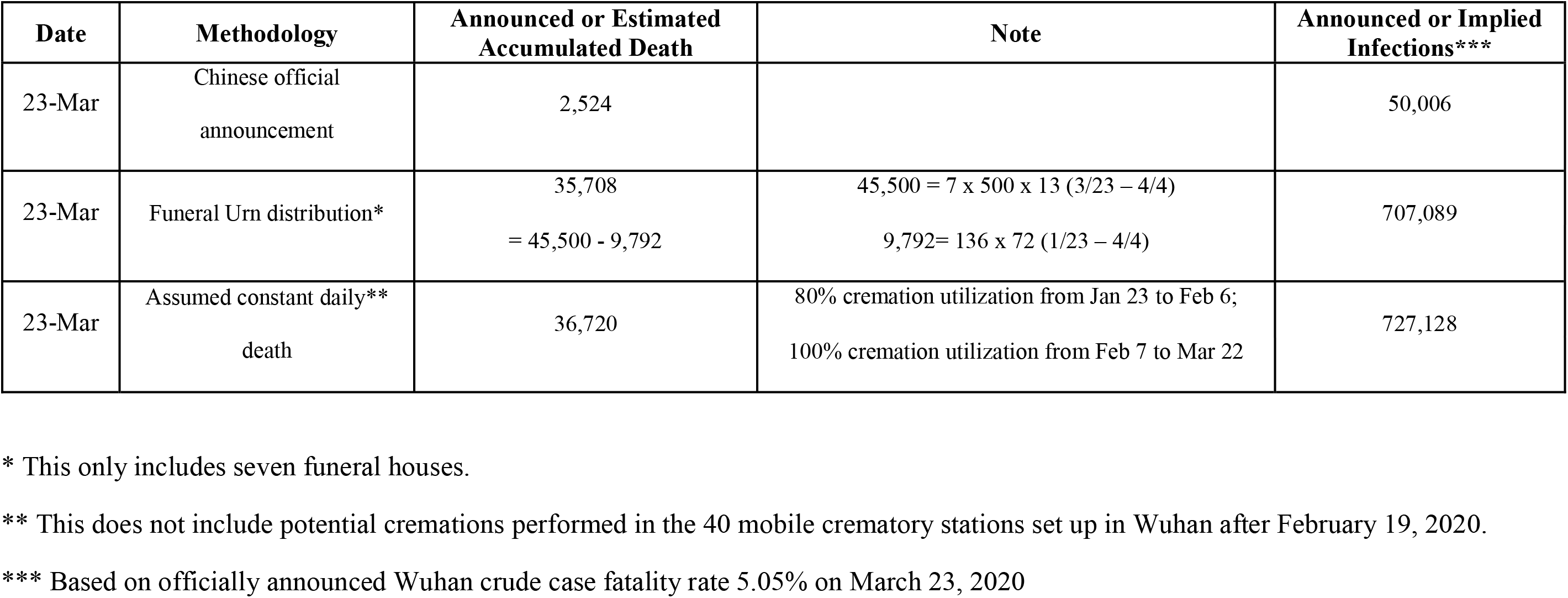
Estimate of COVID-19 Related Deaths in Wuhan, China up to March 23, 2020.

### III. Estimate of COVID related deaths and cumulative infections in all of China

If we use the Wuhan/China ratio based on China’s official data listed in Table 1 (confirmed cases 39.4%), we can extrapolate estimates for China, with cumulative infections potentially reaching 5.67 million on February 7, 2020.

## DISCUSSION

The escalating intervention leading to the unprecedented sudden lockdown of Wuhan and the published medical literature by Chinese researchers and scientists suggest a discrepancy from the officially announced figures of the outbreak. As the epicenter of the COVID-19 outbreak, the necessary epidemiological information about Wuhan was not made available n for the world to initiate plans for a response and to prepare for the potential crisis that is now upon us.

### Challenges for getting needed information in China

A major challenge to an effective response existed in the lack of transparency in reporting by China. With media reports providing fragmented information with no well-defined focus, academic publications subject to selection bias, and possibly less than forthright government data, many challenges have arisen in formulating a data driven approach to this world-wide problem. ^26^ The current study used cremation-related information to estimate epidemiological information including cumulative infections and deaths.

The strength of this approach is that cremation is a common data end point. We applied an exponential growth model during a window period from lockdown until the intervention started to show effects. Note that the approach of the simple exponential growth model is valid under the assumption that countermeasures to slow the spread of the epidemic were ineffective up to the date included. While people claimed that the lockdown in Wuhan combined with the national emergency response have averted 96% of the cases by February 19, 2020,^27^ it should be considered that in the initial period after Wuhan lock down, the effectiveness of countermeasures taken there could be impacted by many factors, including the lack of public awareness, the cross infection at hospitals, the limitation of the medical capacity, the inadequacy of the quarantine facility and space, and the family cluster chain-infections.. However, this is beyond the scope of this manuscript.

A potential weakness of this study is that there is no cremation information in medical literature. All cremation related information came from media reports. To reduce bias, we used media reports from both within China and outside China.^6–12^

### Cumulative infections and deaths from cremation-based analysis

Based on data from the seven of eight Wuhan crematories, our conservative estimates of cumulative infections on February 7 are more than 10 times than those of the official data.

Chinese media raised concerns about infections and deaths beyond the official statistics; for example, when the director of a fever clinic complained that he could admit only five out of 80 potentially infected patients, which suggested a potential 16-fold difference between possible infected patients and confirmed diagnoses in Wuhan. ^4^ The report mentioned the limited nucleic acid-based testing, which was available only after the sequencing data was announced on January 11, 2020. This information also pointed to the limited and exhausted medical facility capacity in Wuhan in late January and early February 2020. In Table 5, we list representative medical facilities in Wuhan and additional support from rest of China. Note that Wuhan was reported to have approximately 95,000 beds by the end of 2018. ^28^ Wuhan designated 100,000 beds for COVID patients by February 20.^29^ This ratio of bed numbers to official confirmed cumulative diagnoses 47,741 as of February 25, 2020 presents a discrepancy which cannot be explained by other causes. Similarly, as of Mar 8, Wuhan had 178,900 health care professionals, and 16,900 ventilators provided to Wuhan, to care for the officially recognized 50,000 (cumulative) patients by that time, is also inexplicable, unless most of the infected were admitted to ICU around the same time.^25^

**Table 5.**
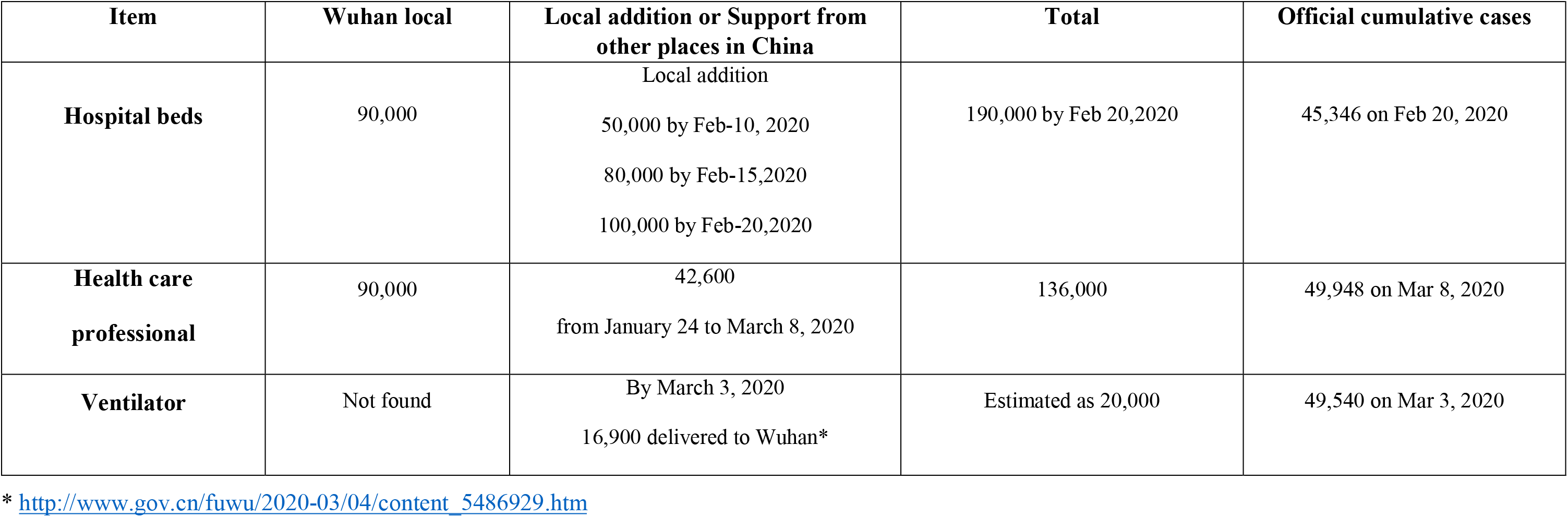
Medical Resources Available in Wuhan after Lockdown (January 23, 2020)

Some researchers have inferred the percentage of undocumented infections, concluding that approximately 86% of all infections were apparently undocumented and that these were the source of infection for 79% of documented cases.^30^ Scissors, examining out-migrants, has also pointed out that China’s COVID-19 figures are arithmetically impossible.^31^ Tsang et. al. estimated that by Feb 20, 2020, there would have been 232,000 (95% CrI (161,000-359,000)) confirmed cases in China as opposed to only 55,508 reported cases.^32^ By using high-resolution domestic travel and infection data, Sanche S et al projected the infected population would be around 233,400 (95% CI 38,757–778,278) by the end of January.^33^ These studies raise similar concerns about the underreporting of Chinese COVID-19 cases.

### Correlation our estimates with funeral urn distribution

Since a major data source is the number of cremations in Wuhan, our estimates have been further verified by the subsequent information of the funeral urn distribution. In late March, *Newsweek* reported that roughly 5,000 urns were shipped to one of the eight Wuhan cremation facilities. The number of urns that arrived in that one facility was already about twice of the city’s official overall death from COVID-19 toll.^34^ Potential COVID-19 related death counts from urn distribution for Wuhan could be 35,708 for seven funeral houses (Table 3). This is consistent with our linear estimate of 36,720, based on the cremation service operation. Both estimates are more than ten times of the official death toll (2,524 on March 23). Additionally, neither took into account those potentially processed in the 40 mobile crematories which were brought to Wuhan after February 19, 2020 for the newly constructed hospitals (Huoshen Shan and Leishen Shan) and “Fangchang” hospitals.^16,17^ The mobile crematory stations can each process up to five tons of “medical waste” including “animal dead bodies” per day. Thus, the calculations here could be significant under-estimates.

### Potential stating time for COVID-19 outbreak in Wuhan

As reported by Huang C et al., symptom onset of the first confirmed case was December 1, 2019.^35^ Under the assumption of 6.4 doubling time, the start dates implied by this study range from October 4, 2019 to October 17, 2019. The estimated “implied start dates” are consistent with other reports where Kristian Andersen suggested a possible “start date” of October 1, 2019 based viral genome analysis.^36^ Under the assumption of 2.54 doubling time, the implied start date is December 14, 2019. However, in this scenario the infection cases were projected to total 2.23 million by February 7, 2020. If the doubling time 2.54 is close to the truth, then China’s official data under reported by millions. If the doubling time 6.4 is close to the truth, there was significant under- and delayed reporting of the COVID-19 epidemic information by China in late January and early February, the critical time for the world to respond and prepare for the pandemic. Given the serious implications of the COVID-19 pandemic, further investigations into this period in China needs to be carried out.

Readers are reminded of the assumptions that underlie our estimates, and they should therefore be taken as approximate. However, even if there were non-negligible reporting errors in these new data, the magnitude of the discrepancy between the results from their analysis and China’s official figures suggests that the potential impact on the global efforts to control the pandemic is obvious. Transparency in China is of critical importance for the world to learn from this infection and for those in the future.

## Data Availability

All needed data are present in the paper, such as the tables and appendix.

---

**Appendix 1.** Eight Crematories/Funeral Houses in Wuhan^6,7^.

| Name of crematory | Number of cremation furnaces | Annual processing (before lockdown) | Daily processing (before lockdown) |
| --- | --- | --- | --- |
| Hankou (汉口殡仪馆) | 30 | 7,000 | 19 |
| Wuchang (武昌殡仪馆) | 18 | 7,000 | 19 |
| Jiangxia (江夏殡仪馆) | 7 | 3,800 | 10 |
| Wuhan Huangpi District<br>(武汉黄陂区殡仪馆) | 7 | 7,000 | 19 |
| Xinzhou District<br>(新洲区殡仪馆) | 5 | 6,000 | 16 |
| Wuhan City Qingshan<br>(武汉市青山殡仪馆) | 7 | 3,700 | 10 |
| Caidian District Yusun Mountain<br>(蔡甸区玉笋山殡仪馆) | 12 | 14,785 | 41 |
| **Total (for seven)** | **86** | **49,285** | **135** |
| Muslim | Information not found | Information not found | Information not found |

**Note:** During normal times, it takes about 2 hours to cremate one body. During lockdown, the process was simplified and it took about one hour to process each body. If seven crematories operate 24 hours nonstop, the maximal number would be $86 \times 24 = 2,064$ .

